# Thyroid hyperplasia and neoplasm adverse events associated with GLP-1 receptor agonists in FDA Adverse Event Reporting System

**DOI:** 10.1101/2023.11.19.23298750

**Authors:** Tigran Makunts, Haroutyun Joulfayan, Ruben Abagyan

## Abstract

Glucagon receptor-like peptide receptor agonists, GLP-1 RAs, are one of the most commonly used drugs for type-2 diabetes mellitus. The clinical guidelines recommend GLP-1 RAs as adjunct to diabetes therapy in patients with chronic kidney disease, presence or risk of atherosclerotic cardiovascular disease, obesity, and other cardiometabolic conditions. The weight loss seen in clinical trials has been explored further in healthy individuals, putting GLP-1 RAs on track to be the next weight loss treatment. Although the adverse event profile is relatively safe, most GLP-1 RAs come with a labeled black boxed warning of the risk of thyroid cancers, based on animal models and some postmarketing case reports in humans. Considering the increasing popularity of this drug class and its expansion into a new popular indication, a further review of most recent postmarketing safety data is warranted to quantify thyroid hyperplasia and neoplasms instances. In this study we analyzed over eighteen million reports from United States Food and Drug Administration Adverse Event Reporting System and identified 17,653 relevant GLP-1 RA monotherapy reports to provide the evidence of significantly increased propensity for thyroid hyperplasias and neoplasms in patients taking GLP-1 RA as monotherapy when compared to patients taking sodium-glucose cotransporter-2 inhibitor monotherapy.

## Introduction

Glucagon receptor-like peptide receptor agonists (GLP-1 RAs) have gained increased popularity due to improved safety and efficacy profiles observed in clinical. This class of drugs includes liraglutide, semaglutide, exenatide, and dulaglutide^1-5^. GLP-1 RAs are indicated as a therapeutics adjunct to diet and exercise, to improve glycemic control in patients with type-2 diabetes mellitus (T2DM). The American Association of Clinical Endocrinology and the American Diabetes Association recommend GLP-1 RA for T2DM patients with chronic kidney disease, or atherosclerotic cardiovascular disease risk, or obesity^6,7^. GLP-1 RAs have also been utilized in cardiometabolic conditions of wide range, including but not limited to non-alcoholic fatty liver disease, non-alcoholic steatohepatitis^8^.

GLP-1 RAs were recommended for obese T2DM patients because of the weight loss observed during the clinical trials^2-5,9^. The effect was attributed to decreasing gastric emptying, peristalsis, appetite, and glucose absorption^10,11^. This effect was further explored in studies of non-T2DM populations^12-14^, suggesting the expansion of GLP-1 RA use.

Common adverse events associated with GLP-1 RAs observed during the clinical trials include nausea, hypoglycemia, vomiting, diarrhea, feeling jittery, dizziness, headache, and dyspepsia. Of a greater concern are the labeled Black Boxed Warnings of liraglutide, semaglutide, and dulaglutide, marking these as contraindicated in patients with family history of medullary thyroid carcinoma. These warnings were based on non-human data about the development of thyroid C-cell tumors in rats and mice receiving clinically relevant doses of GLP-1 RAs^15-17^. The thyroid cancer association in humans has been studied and observed as well in retrospective studies^18-20^. However, there is conflicting evidence from a meta-analysis of human randomized controlled trials which refutes this association^21^.

The increased popularity of GLP-1 RAs and the unsettled association of thyroid hyperplasias and neoplasms prompted further investigation into the most recent FDA Adverse Event Reporting System (FAERS) data sets. In this study we evaluated thyroid hyperplasia and neoplasm related adverse events reported as associated with GLP-1 RA monotherapy use when compared to Sodium-glucose Cotransporter-2 RAs (SGLT-2) inhibitors. We analyzed GLP-1 RAs individually and as a class.

## Methods

### FDA Adverse Event Reporting System

The FDA Adverse Event Reporting System is a repository of AE cases sent to the FDA through MedWatch (form 3500/3500a)^22,23^. The cases include AEs submitted voluntarily by healthcare professionals, individuals, legal representatives, and spontaneous mandatory reports by manufacturers/sponsors. At the time of the analysis FAERS/AERS contained 18,274,795 reports from January 2004 to September 2022.

The data sets have been de-identified and made available online at: https://fis.fda.gov/extensions/FPD-QDE-FAERS/FPD-QDE-FAERS.html

### Data preparation, cohort selection, and outcome measure

The FAERS quarterly data sets were initially downloaded in text format. Due to variability of data structure between quarters and paucity of some of the variables, the cases were standardized to fit a uniform structure^24-26^.

Out of the total of 18,274,795 reports, AE cases where the report was submitted by a healthcare professional (pharmacists, physicians, nurses, other healthcare professionals) were selected into the initial data set (n=6,360,489). This allowed for minimizing reporting bias and adding to the clinical relevance of the studied cases. Further, reports where GLP-1 RAs were the only drug reported (further referred to as *monotherapy*) were selected into the GLP-1 RA cohort (n=17,653) to avoid potential confounding effects from concomitant medications. The cohort was further split into individual GLP-1 RA sub-cohorts: semaglutide (n=3,230), dulaglutide (n=3,768), exenatide (n=4,493), liraglutide (n=6,162). SGLT-2 inhibitor monotherapy reports (canagliflozin, dapagliflozin, and empagliflozin, n=14,102) were selected as a control cohort. SGLT-2 inhibitors were selected as a control due to the comparable recommendation by diabetes management guidelines. Metformin monotherapy was initially selected as the control cohort to match the population of the cohort of interest (n=8,536) however only a single case of interest (Preferred Term [PT] code: thyroid cancer) was reported. The undetectable baseline of this AE made it impossible to use metformin as a control (Table 1). All the thyroid related AE PT codes based on standard MedDRA queries (SMQs) and FDA Medical queries (FMQs) were used in the case selection process. Disproportionality analysis using reporting odds ratios (ROR) and 95% confidence intervals were used for determination of the statistical significance of the results.

**Table 1.**
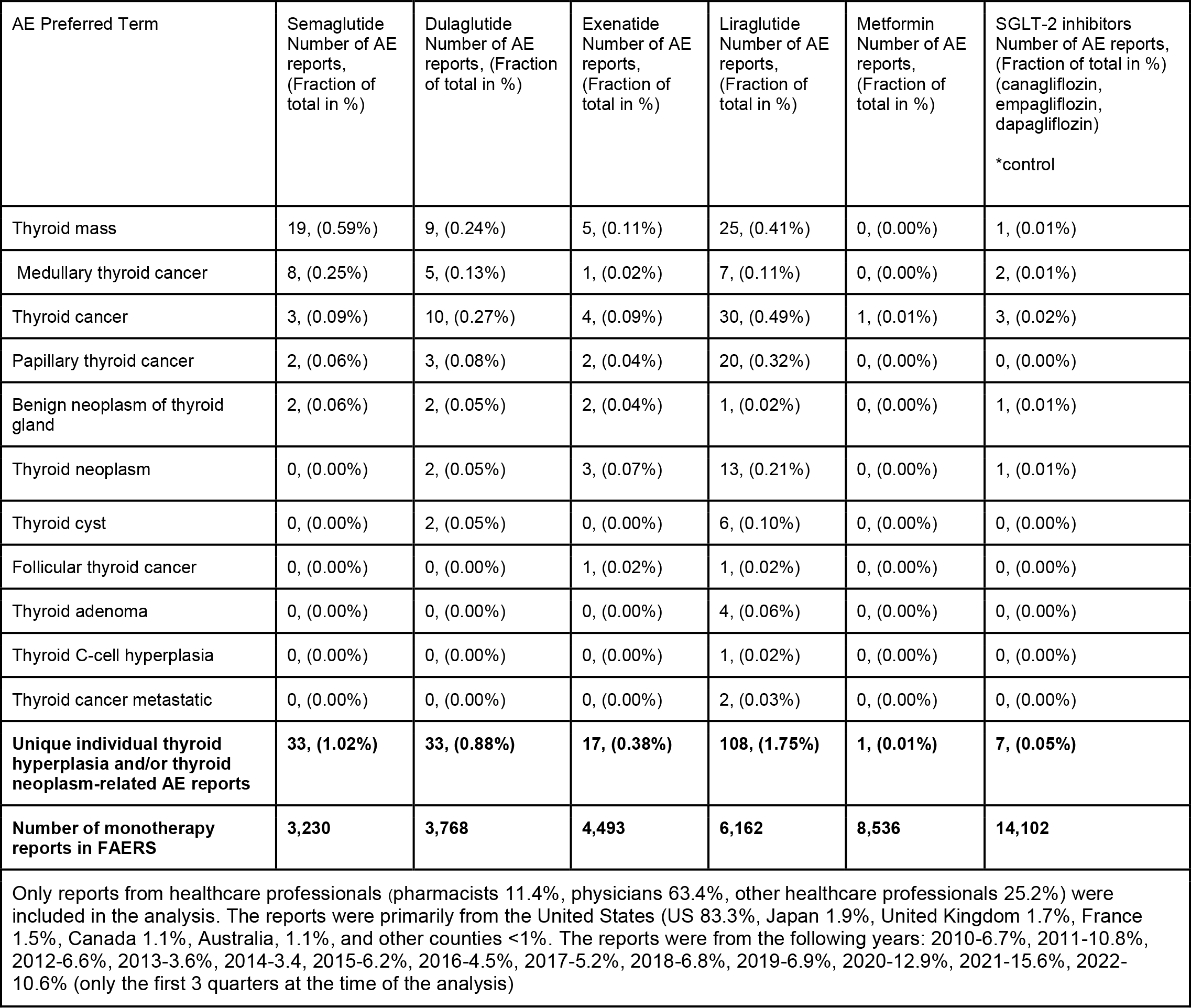
Thyroid hyperplasia and/or thyroid neoplasm-related AE reports in GLP-1 RA, metformin and SGLT1 FAERS/AERS reports.

### Statistical Analysis

#### Descriptive statistics

Frequencies for each AE PT code were calculated by the equation:

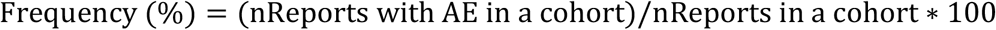

#### Comparative Statistics

AE report rates were compared via the Reporting Odds Ratio (ROR) analysis using the following equations:

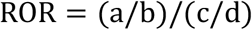

where

Number of AE cases in exposed group

Number of no AE cases in exposed group

Number of AE cases in control group

Number of no AE cases in control group

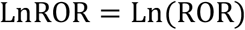

Standard Error of Log Reporting Odds Ratio;

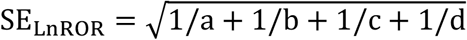

95% Confidence Interval;

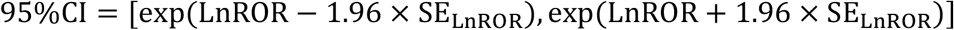

## Results

A total of 31,755 monotherapy reports, including 17,653 GLP-1 RA and 14,102 SGLT-2 inhibitor reports, were used for the analysis of 191 unique thyroid hyperplasia/neoplasm reports in the GLP-1 RA group, and 7 reports in the SGLT-2 group respectively (Tables 1 and 2).

**Table 2.** AE report number and respective demographics.

| Drug cohort | Semaglutide | Dulaglutide | Exenatide | Liraglutide | SGLT-2 inhibitors<br>*control |
| --- | --- | --- | --- | --- | --- |
| Total (unique individual hyperplasia/neoplasm reports) | 33, (1.02) | 33, (0.88) | 17, (0.38) | 108, (1.75) | 7, (0.05) |
| Sex | M: 4<br>F: 27<br>Unk: 2 | M: 14<br>F: 14<br>Unk: 5 | M: 3<br>F: 13<br>Unk: 1 | M: 35<br>F: 66<br>Unk: 7 | M: 3<br>F: 2<br>Unk: 2 |
| Age | Median: 56.0<br>Mean: 55.94<br>St. dev: 13.41 | Median: 60<br>Mean: 57.33<br>St. dev: 11.35 | Median: 61.0<br>Mean: 64.67<br>St. dev: 6.35 | Median: 55<br>Mean: 53.07<br>St. dev: 11.08 | (no age reports)<br>Median: -<br>Mean: -<br>St. dev: - |

GLP-1 RAs monotherapy reports manifested a statistically significant increase in thyroid hyperplasia/neoplasm AEs, reported odds ratio (ROR) being 21.80, 95% (10.25, 46.36) when compared to SGLT-2 inhibitors. When analyzed individually, every GLP-1 RA had significantly increased reporting of those thyroid AEs when compared to SGLT-2 inhibitors. Results for all the GLP-1 RAs show the lower bound of the 95% CI of the ROR range to be above 3: semaglutide-ROR 20.58, (95%CI [9.19, 46.57]), dulaglutide-ROR 17.6 (95%CI [7.80, 39.92]), exenatide-ROR 7.62 (95%CI [3.16, 18.39]), and liraglutide-ROR 35.31 (95%CI [16.43, 75.88]) (Fig 1).

**Fig. 1.**
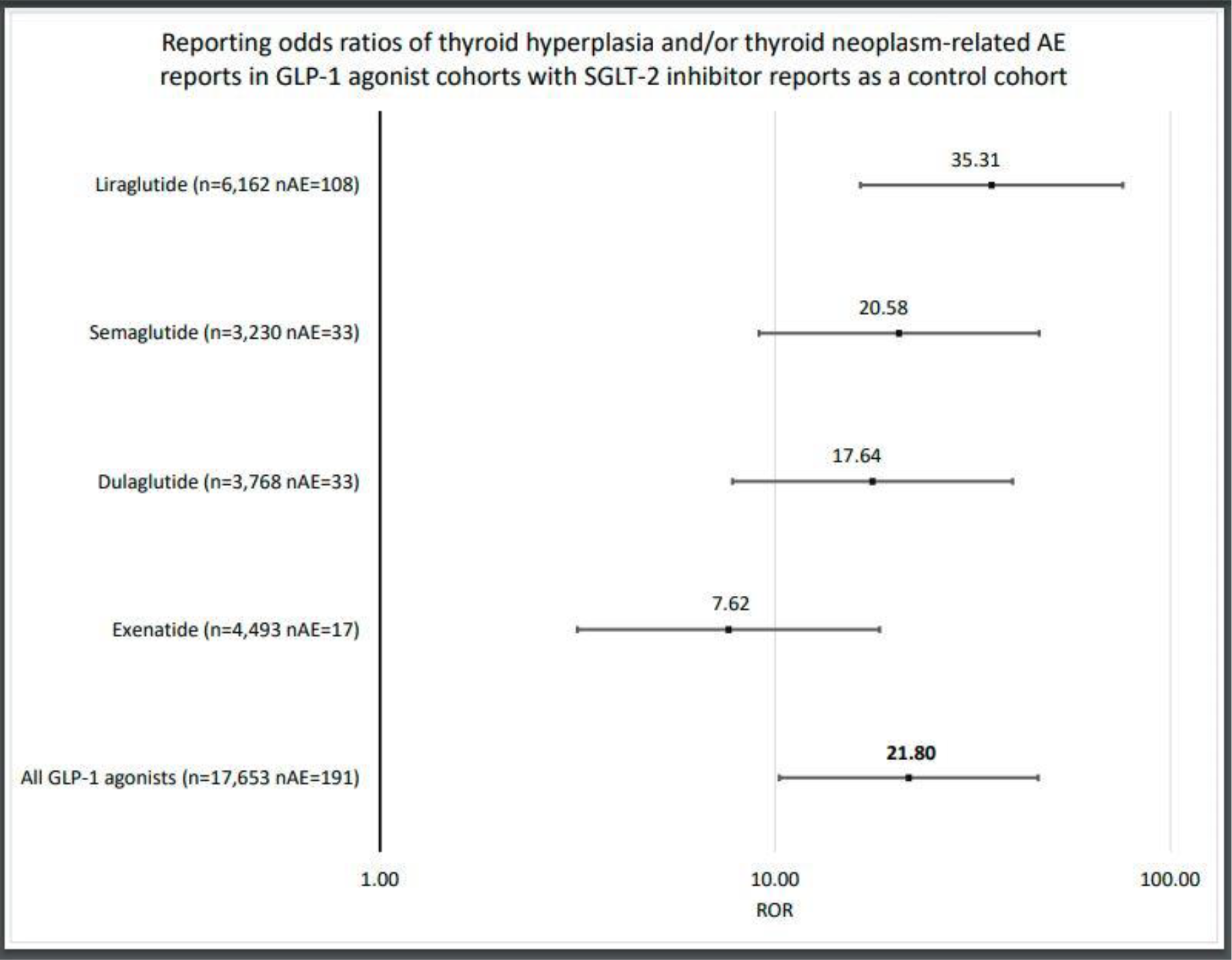
Reporting odds ratios of thyroid hyperplasia/neoplasm AEs in GLP-1 RA, as a class, monotherapy cohort, and in individual GLP-1 RA cohorts, when compared to SGLT-2 inhibitor monotherapy control. X-axis presented in logarithmic scale.

### Discussion

In this study we observed a reported association of GLP-1 RAs treatment with thyroid hyperplasia and neoplasm AEs. To our knowledge, this is the first analysis of the FDA Adverse Event Reporting System (FAERS) to generate a reported odds ratio profile of GLP-1 RA monotherapy as a class and further analyze the individual GLP-1 RA monotherapy AEs compared to SGLT-2 inhibitors monotherapy ones. The mean age of the specific drug cohorts with these AEs was close for comparison, ranging from 53.0 to 64.7 years. The thyroid hyperplasia and neoplasm AE association was significant for every GLP-1 RAs in the class, with an even narrower range of 95% CI in the combined cohort. The RORs ranged from 7.62 to 35.31, with the lowest bound being 7.80. The highest mean ROR value was observed for liraglutide, and the lowest mean ROR was observed for exenatide, with only a small overlap in the 95% confidence intervals. Interestingly, this trend was also seen in a similar analysis performed by Mali et al.^20^ using European pharmacovigilance database (EudraVigilance) where this association was strongest in the liraglutide cohort followed by the exenatide cohort with proportional reporting ranges of 27.5 (95% CI, 22.7-33.3) and 22.5 (95% CI, 17.9-28.3), respectively. The differences in the numbers and CI ranges are due to the numbers of reports and type of analysis.

An association between T2DM and thyroid function has been previously established^27^. These disease states are often comorbid, and one affects the disease progression of the other^28^. However, the potential molecular mechanisms responsible for thyroid effects of GLP-1 RAs are insufficiently characterized and may include multiple pathways such as phosphoinositol-3 kinase/AKT serine/threonine kinase, mitogen-activated protein kinase/extracellular signal-regulated kinase pathways, expression GLP-1 receptors by C-cells, and GLP-1 association with triiodothyronine levels^21,29^. Thyroid hormone plays a pivotal role in nearly every aspect of lipid metabolism^30,31^. Thus, it was expected to observe thyroid hyperplasia and neoplasm related AEs across all the T2DM drug cohorts that were investigated. However, only a single report of thyroid hyperplasia/neoplasm AE (thyroid cancer) was observed in metformin monotherapy cohort, and only seven were observed in the SGLT-2 monotherapy cohort which was selected as control due to similar T2DM disease stage treatment guideline recommendation. In contrast, almost 200 of these AEs were reported for GLP-1 RAs in similarly sized cohorts, resulting in statistically significant reported association. Therefore, as the GLP-1 receptor agonists expand into non-T2DM indications such as obesity metabolic syndrome and other related conditions, controlled studies and better understanding of the molecular mechanisms of action are necessary to investigate this association and prevent potential serious consequences.

### Study limitations

Since this is an association study, the causality between the thyroid hyperplasias/neoplasms and GLP-1 RAs was not clinically adjudicated. However, this analysis of a large population scale AE database provides a strong signal that may be clinically significant. The numbers of AEs presented in the study do not represent all treated patients due to voluntary submissions resulting in over- and under-reporting^32,33^. Additionally, due the nature and long progression of the studied AEs, often requiring invasive procedures for a proper diagnosis, the cases may be significantly underreported. The case narrative with additional information such as thorough medical history and test/diagnostic results were not available in the data sets provided by the FDA. Other limitations include the possible over the counter medications and supplements that are often not reported to healthcare professionals and may potentially add noise to the cohort compositions, AE frequencies and, to a lesser extent, reported odds ratios and respective confidence intervals. However, considering the large number of individuals in the GLP-1 RA and the control cohorts, matched by indication, the signal was statistically significant.

The case narratives are not included in the FAERS database due to privacy concerns.

## Data Availability

Data Availability Statement
The datasets generated and/or analyzed during the current study are available in the FAERS repository, https://fis.fda.gov/extensions/FPD-QDE-FAERS/FPD-QDE-FAERS.html

https://fis.fda.gov/extensions/FPD-QDE-FAERS/FPD-QDE-FAERS.html

## Acknowledgements

We thank Dr. Vahe Melkonyan, MD, for helpful discussions. We also that Dr. Da Shi and Chris Edwards for help with data processing.

## Author information

### Authors and Affiliations

Skaggs School of Pharmacy and Pharmaceutical Sciences, University of California San Diego, La Jolla, CA, USA

Tigran Makunts, Haroutyun Joulfayan & Ruben Abagyan

### Contributions

H.J. performed the experiments, R.A. and T.M. designed the study and, R.A., H.J, and T.M. drafted the manuscript and reviewed the final version. R.A. processed the data set. This work was funded in part by NIH R35GM131881.

### Corresponding author

Correspondence to Ruben Abagyan.

## Ethics declarations

### Ethics Statement

Ethical review and approval was not required for the study on human participants in accordance with the local legislation and institutional requirements. Written informed consent from the participants’ legal guardian/next of kin was not required to participate in this study in accordance with the national legislation and the institutional requirements.

## Data Availability Statement

The datasets generated and/or analyzed during the current study are available in the FAERS repository, https://fis.fda.gov/extensions/FPD-QDE-FAERS/FPD-QDE-FAERS.html

## References

1. Ahmann AJ, Capehorn M, Charpentier G, et al. Efficacy and Safety of Once-Weekly Semaglutide Versus Exenatide ER in Subjects With Type 2 Diabetes (SUSTAIN 3): A 56-Week, Open-Label, Randomized Clinical Trial. Diabetes Care. Feb 2018;41(2):258–266. doi:10.2337/dc17-0417

2. Sorli C, Harashima SI, Tsoukas GM, et al. Efficacy and safety of once-weekly semaglutide monotherapy versus placebo in patients with type 2 diabetes (SUSTAIN 1): a double-blind, randomised, placebo-controlled, parallel-group, multinational, multicentre phase 3a trial. Lancet Diabetes Endocrinol. Apr 2017;5(4):251–260. doi:10.1016/S2213-8587(17)30013-X

3. Wysham C, Blevins T, Arakaki R, et al. Efficacy and safety of dulaglutide added onto pioglitazone and metformin versus exenatide in type 2 diabetes in a randomized controlled trial (AWARD-1). Diabetes Care. Aug 2014;37(8):2159–67. doi:10.2337/dc13-2760

4. Garber A, Henry R, Ratner R, et al. Liraglutide versus glimepiride monotherapy for type 2 diabetes (LEAD-3 Mono): a randomised, 52-week, phase III, double-blind, parallel-treatment trial. Lancet. Feb 07 2009;373(9662):473–81. doi:10.1016/S0140-6736(08)61246-5

5. Buse JB, Bergenstal RM, Glass LC, et al. Use of twice-daily exenatide in Basal insulin-treated patients with type 2 diabetes: a randomized, controlled trial. Ann Intern Med. Jan 18 2011;154(2):103–12. doi:10.7326/0003-4819-154-2-201101180-00300

6. Garber AJ, Abrahamson MJ, Barzilay JI, et al. CONSENSUS STATEMENT BY THE AMERICAN ASSOCIATION OF CLINICAL ENDOCRINOLOGISTS AND AMERICAN COLLEGE OF ENDOCRINOLOGY ON THE COMPREHENSIVE TYPE 2 DIABETES MANAGEMENT ALGORITHM--2016 EXECUTIVE SUMMARY. Endocr Pract. Jan 2016;22(1):84–113. doi:10.4158/EP151126.CS

7. Blonde L, Umpierrez GE, Reddy SS, et al. American Association of Clinical Endocrinology Clinical Practice Guideline: Developing a Diabetes Mellitus Comprehensive Care Plan-2022 Update. Endocr Pract. Oct 2022;28(10):923–1049. doi:10.1016/j.eprac.2022.08.002

8. Muzurović EM, Volčanšek Š, Tomšić KZ, et al. Glucagon-Like Peptide-1 Receptor Agonists and Dual Glucose-Dependent Insulinotropic Polypeptide/Glucagon-Like Peptide-1 Receptor Agonists in the Treatment of Obesity/Metabolic Syndrome, Prediabetes/Diabetes and Non-Alcoholic Fatty Liver Disease-Current Evidence. J Cardiovasc Pharmacol Ther. 2022;27:10742484221146371. doi:10.1177/10742484221146371

9. Vilsbøll T, Christensen M, Junker AE, Knop FK, Gluud LL. Effects of glucagon-like peptide-1 receptor agonists on weight loss: systematic review and meta-analyses of randomised controlled trials. BMJ. Jan 10 2012;344:d7771. doi:10.1136/bmj.d7771

10. Shaefer CF, Kushner P, Aguilar R. User’s guide to mechanism of action and clinical use of GLP-1 receptor agonists. Postgrad Med. 2015;127(8):818–26. doi:10.1080/00325481.2015.1090295

11. Drucker DJ. GLP-1 physiology informs the pharmacotherapy of obesity. Mol Metab. Mar 2022;57:101351. doi:10.1016/j.molmet.2021.101351

12. Zhang F, Tong Y, Su N, et al. Weight loss effect of glucagon-like peptide-1 mimetics on obese/overweight adults without diabetes: A systematic review and meta-analysis of randomized controlled trials. J Diabetes. May 2015;7(3):329–39. doi:10.1111/1753-0407.12198

13. Rubino DM, Greenway FL, Khalid U, et al. Effect of Weekly Subcutaneous Semaglutide vs Daily Liraglutide on Body Weight in Adults With Overweight or Obesity Without Diabetes: The STEP 8 Randomized Clinical Trial. JAMA. Jan 11 2022;327(2):138–150. doi:10.1001/jama.2021.23619

14. Dushay J, Gao C, Gopalakrishnan GS, et al. Short-term exenatide treatment leads to significant weight loss in a subset of obese women without diabetes. Diabetes Care. Jan 2012;35(1):4–11. doi:10.2337/dc11-0931

15. Bjerre Knudsen L, Madsen LW, Andersen S, et al. Glucagon-like Peptide-1 receptor agonists activate rodent thyroid C-cells causing calcitonin release and C-cell proliferation. Endocrinology. Apr 2010;151(4):1473–86. doi:10.1210/en.2009-1272

16. Madsen LW, Knauf JA, Gotfredsen C, et al. GLP-1 receptor agonists and the thyroid: C-cell effects in mice are mediated via the GLP-1 receptor and not associated with RET activation. Endocrinology. Mar 2012;153(3):1538–47. doi:10.1210/en.2011-1864

17. Byrd RA, Sorden SD, Ryan T, et al. Chronic Toxicity and Carcinogenicity Studies of the Long-Acting GLP-1 Receptor Agonist Dulaglutide in Rodents. Endocrinology. Jul 2015;156(7):2417–28. doi:10.1210/en.2014-1722

18. Bezin J, Gouverneur A, Pénichon M, et al. GLP-1 Receptor Agonists and the Risk of Thyroid Cancer. Diabetes Care. Nov 10 2022;doi:10.2337/dc22-1148

19. Yang Z, Lv Y, Yu M, et al. GLP-1 receptor agonist-associated tumor adverse events: A real-world study from 2004 to 2021 based on FAERS. Front Pharmacol. 2022;13:925377. doi:10.3389/fphar.2022.925377

20. Mali G, Ahuja V, Dubey K. Glucagon-like peptide-1 analogues and thyroid cancer: An analysis of cases reported in the European pharmacovigilance database. J Clin Pharm Ther. Feb 2021;46(1):99–105. doi:10.1111/jcpt.13259

21. Hu W, Song R, Cheng R, et al. Use of GLP-1 Receptor Agonists and Occurrence of Thyroid Disorders: a Meta-Analysis of Randomized Controlled Trials. Front Endocrinol (Lausanne). 2022;13:927859. doi:10.3389/fendo.2022.927859

22. Craigle V. MedWatch: The FDA Safety Information and Adverse Event Reporting Program. J Med Libr Assoc. 2007;95(2)(2):224–225. doi:10.3163/1536-5050.95.2.224

23. Kessler DA. Introducing MEDWatch. A new approach to reporting medication and device adverse effects and product problems. JAMA. Jun 02 1993;269(21):2765–8. doi:10.1001/jama.269.21.2765

24. Keshishi D, Makunts T, Abagyan R. Common osteoporosis drug associated with increased rates of depression and anxiety. Sci Rep. Dec 14 2021;11(1):23956. doi:10.1038/s41598-021-03214-x

25. Wollmer MA, Makunts T, Krüger THC, Abagyan R. Postmarketing safety surveillance data reveals protective effects of botulinum toxin injections against incident anxiety. Sci Rep. 12 21 2021;11(1):24173. doi:10.1038/s41598-021-03713-x

26. Makunts T, Jerome L, Abagyan R, de Boer A. Reported Cases of Serotonin Syndrome in MDMA Users in FAERS Database. Front Psychiatry. 2021;12:824288. doi:10.3389/fpsyt.2021.824288

27. Kadiyala R, Peter R, Okosieme OE. Thyroid dysfunction in patients with diabetes: clinical implications and screening strategies. Int J Clin Pract. Jul 2010;64(8):1130–9. doi:10.1111/j.1742-1241.2010.02376.x

28. Jali MV, Kambar S, Jali SM, Pawar N, Nalawade P. Prevalence of thyroid dysfunction among type 2 diabetes mellitus patients. Diabetes Metab Syndr. Nov 2017;11 Suppl 1:S105–S108. doi:10.1016/j.dsx.2016.12.017

29. Waser B, Blank A, Karamitopoulou E, Perren A, Reubi JC. Glucagon-like-peptide-1 receptor expression in normal and diseased human thyroid and pancreas. Mod Pathol. Mar 2015;28(3):391–402. doi:10.1038/modpathol.2014.113

30. Duntas LH, Brenta G. A Renewed Focus on the Association Between Thyroid Hormones and Lipid Metabolism. Front Endocrinol (Lausanne). 2018;9:511. doi:10.3389/fendo.2018.00511

31. Pucci E, Chiovato L, Pinchera A. Thyroid and lipid metabolism. Int J Obes Relat Metab Disord. Jun 2000;24 Suppl 2:S109–12. doi:10.1038/sj.ijo.0801292

32. Alatawi YM, Hansen RA. Empirical estimation of under-reporting in the U.S. Food and Drug Administration Adverse Event Reporting System (FAERS). Expert Opin Drug Saf. Jul 2017;16(7):761–767. doi:10.1080/14740338.2017.1323867

33. Maciejewski M, Lounkine E, Whitebread S, et al. Reverse translation of adverse event reports paves the way for de-risking preclinical off-targets. Elife. Aug 2017;6 doi:10.7554/eLife.25818

